# Serum levels of the novel adipokine isthmin-1 are associated with obesity in pubertal boys

**DOI:** 10.1101/2022.03.02.22271664

**Authors:** Francisco Javier Ruiz-Ojeda, Augusto Anguita-Ruiz, Maria C. Rico, Rosaura Leis, Gloria Bueno, Mercedes Gil-Campos, Ángel Gil, Concepción M Aguilera

## Abstract

**Purpose:** The aims of this study were; 1) to evaluate if there is an association between the serum levels of the novel insulin-like adipokine isthmin-1 (ISM1) and obesity-related phenotypes in a population of Spanish children, 2) to investigate the plausible molecular alterations behind the alteration of the serum levels of this protein in children with obesity.

**Methods:** The study population is a sub-cohort of the PUBMEP research project, consisting of a cross-sectional population of 119 pubertal children with overweight (17 boys, 19 girls), obesity (20 boys, 25 girls) and normal weight (17 boys, 21 girls). All subjects were classified into experimental groups according to their sex, obesity and insulin resistance (IR) status. They counted on anthropometry, glucose, and lipid metabolism, inflammation and cardiovascular biomarkers as well as ISM1 serum levels measured. This population was intended as a discovery population in which to elucidate the relationship between obesity and ISM1 levels in children. Furthermore, the study population had blood whole-genome DNA methylation allowing deepening into the obesity-ISM1 molecular relationship.

**Results:** Higher serum levels of ISM1 were observed in boys with obesity when compared with normal-weight (P=0.004), and overweight (P=0.007). ISM1 serum levels were positively associated with BMI *Z-*score (P=0.005), and negatively with myeloperoxidase (MPO) (p=0.043) in boys. Nevertheless, we did not find associations between ISM1 serum levels and metabolic outcomes in girls, indicating a putative sexual dimorphism. DNA methylation levels in two-enhancer-related CpG sites of ISM1 (cg03304641 and cg14269097) were associated with serum levels of ISM1 in children.

**Discussion:** We report an unprecedented study that provides a major step forward showing that ISM1 is robustly associated with obesity in pubertal boys, elucidating how this protein might be of special relevance as a new biomarker of obesity in children.

## Implications and Contribution

Study findings reveal differences in the circulating levels of the novel insulin-like adipokine ISM1 between pubertal children with obesity and normal-weight, and different associations with metabolic traits, which may be further investigated to be used as a potential biomarker of obesity in young populations.

Childhood obesity is increasing globally and total adiposity is the key driver of metabolic risk in children and adolescent, which represent a strong risk factor for insulin resistance (IR) and future type 2 diabetes (T2D) [1]. Puberty is a time of metabolic and hormonal changes, and it is associated with a reduced insulin sensitivity that recovers at puberty completion in only some children [2-4]. In growing children, adipocyte hypertrophy is associated with inflammation and local and systemic IR, independently of BMI and fat mass, being adipose tissue essential to maintain a functional metabolism [5]. However, the molecular mechanism of IR is still unknown, particularly in children. This is in part because processes such as growth and puberty affect insulin secretion and sensitivity [6]. Hence, understanding the molecular and biological processes underlying metabolic changes (glucose and lipid regulation) during puberty, and identifying pathways and biomarkers that might help to increase peripheral glucose uptake would be beneficial to reduce the impact of obesity and to prevent T2D. A recent paper reported that the protein isthmin-1 (ISM1) is secreted by mature adipocytes and triggers a signaling cascade similar to that of insulin. The novel adipokine acts through a unidentified receptor tyrosine kinase and, at pharmacological doses in mice, ISM1 ameliorates metabolic disturbances associated with T2D, including hyperglycemia and liver steatosis [7, 8]. Here, we report that serum levels of the novel insulin-like adipokine ISM1 is, indeed, associated with obesity in pubertal boys, but not in girls in a well-characterized population of Spanish children under a cross-sectional design. Furthermore, we identified DNA methylation in two-enhancer-related CpG sites of the ISM1 region (cg14269097 and cg14269097) associated with serum levels of ISM1 in children with obesity.

## Methods

This study was conducted within the context of the multicenter PUBMEP study in Spain “Puberty and metabolic risk in children with obesity”, previously published [9, 10]. Here, a sub-population of 119 pubertal children (54 boys and 65 girls) from the whole PUBMEP cohort was selected for analysis. A total of 38 were normal-weight (17 boys), 36 overweight (17 boys) and 45 children with obesity (20 boys). The following characteristics were considered as exclusion criteria: birth weight <2500 g; intake of any drug that could alter blood glucose, blood pressure or lipid metabolism; not being able to comply with the study procedures and being participating or having participated in the last three months in an investigation project. This study was conducted according to the guidelines set out in the Declaration of Helsinki (Edinburgh 2000 revised), and all procedures were approved by the Ethics and Research Committee of Galicia Autonomous Community (2011/198 and 2016/522). Written consent was obtained from the parents of all the children. Anthropometric measurements such as body weight (kg), height (cm), hip circumference (cm) and waist circumference (WC) (cm) were measured using standardized procedures. BMI z-score was calculated based on the Spanish reference standards published. Blood pressure was measured three times for each individual by the same examiner using a mercury sphygmomanometer and following international recommendations [11]. Measures of lipid and glucose metabolism, hormones and classical biochemical parameters were performed at the laboratories of each participating hospital following internationally accepted quality control protocols. Blood samples were collected in overnight fasting conditions, centrifuged, and plasma and serum were stored at -80°C. The presence of IR in children was defined according to the HOMA insulin resistance (HOMA-IR) index. The cut-off points were obtained from a previously well-described Spanish cohort composed of children and adolescents [12, 13]. The cut-off points for IR were based on the 95^th^ HOMA-IR percentile, considering sex (HOMA-IR ≥ 3.38 in boys and HOMA-IR ≥ 3.90 in girls). These cut-off points have already been tested and validated as good metabolic risk classifiers in our population according to the results from a previous PUBMEP report [10]. Plasma adipokines, inflammation, and cardiovascular risk biomarkers (adiponectin, leptin, resistin, tumor necrosis factor alpha (TNF-α), high-sensitivity CRP (hsCRP), interleukin (IL)-6, IL-8, total plasminogen activator inhibitor-1 (PAI-1), P-selectin, myeloperoxidase (MPO), monocyte chemoattractant protein 1 (MCP-1), soluble intercellular cell adhesion molecule-1 sICAM-1, and soluble vascular cell adhesion molecule-1 (sVCAM)) were analyzed in all samples and time points using XMap technology (Luminex Corporation, Austin, TX) and human monoclonal antibodies (Milliplex Map Kit; Millipore, Billerica, MA) as previously reported [10, 14]. Descriptive data are expressed as mean (standard deviation) or median [min-max] if not normally distributed. One-way ANOVA, Kruskal-Wallis and the Welch test were employed to assess group differences. DNA methylation analysis was carried out by using the Infinium MethylationEPIC microarray using bead chip technology (Illumina, San Diego, CA, USA) as previously described [10]. ISM1 protein levels were determined in serum using the Human SEQ515Hu for ISM 1 (Cloud-Clone Corp., USA), an enzyme-linked immune-absorbent assay kit according to the manufacturers’ instructions. The coefficient of variance was 4%. Two-way ANOVA and Tukey’s multiple comparisons test were employed to assess groups’ differences in ISM1 levels between boys and girls and normal-weight, overweight and obese children. Multiple linear regression (MLR) analyses were applied for all continuous variables to study their association with ISM1 levels. In these analyses, origin, age, gender, pubertal stage, height, BMI *Z*-score, and insulin were adjusted as confounders when necessary. A P-value <0.05 was considered as significant. Given the number of analyzed outcomes, we considered false discovery rate (FDR) as in Benjamini and Hochberg to correct for multiple hypothesis testing. MLRs were also applied for all calculated deltas to study their correlation with the change in ISM1 levels. All described analyses were performed in R environment version 3.6.0.

## Results

General characteristics of the 119 children in this cross-sectional study are shown in **Supplementary Table 1**. ISM1 serum levels according to obesity by sex are shown in **Fig. 1A**. Higher ISM1 serum levels were observed in boys with obesity when compared with normal-weight (P = 0.0037) and overweight (P=0.0071), non-adjusted. However, no changes were observed in girls. When all subjects of the sample were compared together, we found higher levels in children with obesity compared with those with normal weight (P = 0.0413) and with those overweight (P = 0.0102). A trend to increase the ISM1 levels in boys with IR was observed, but no statistical differences were shown between normal-weight non-IR, non-IR and IR children (**Fig. 1B**). To elucidate the relationship between ISM1 and obesity, MLRs were further conducted in a wide range of metabolic outcomes separated by sex (**Tables 1-2**) and properly adjusted by confounders such as age, origin, height, BMI and IR when applicable. In boys, the strongest association was found for BMI *Z*-score (P=0.005), but not in girls (**Fig. 2A and S1A**). MPO (P=0.043) was negatively associated with ISM1 serum levels in boys. All of them were properly adjusted for confounders (please, see **Table 1 and 2** legends), both in boys and girls. Surprisingly, though both ISM1 and leptin levels are significantly higher in children with obesity, no significant associations were identified with leptin neither in boys nor in girls (**Fig. 2B**). Fifty-one methylation sites were selected from the Infinium Methylation EPIC microarray of which two were annotated as promoter-associated CpGs (**Supplementary Table 2**). All the CpGs were annotated as open sea. We found a positive significant association between the methylation status of the probe cg03304641 and ISM1 serum levels in pubertal children (P=0.0062) (**Fig. 3A, S2A and Supplementary Table 2**). However, for the probe cg14269097 there was a negative association with ISM1 serum levels (P=0.038) (**Fig. 3B, S2B and Supplementary Table 2**).

**Table 1.** Association between the change in ISM1 serum levels and the change in metabolic outcomes in boys in the cross-sectional population (PUBMEP study). Multiple regression analyses with the change in ISM1 serum levels as independent variable. Models were adjusted for the change in BMI *Z*-Score, the origin, and the pubertal stage reached. When the dependent variable was the change in the BMI *Z*-Score, we replaced the BMI confounder with the change in Insulin levels. The change in Height was further included in models when the dependent variable was change in Blood pressure. Abbreviations: SE, standard error; CI, confidence interval; AST, aspartate aminotransferase; ALT, alanine aminotransferase; BMI, body mass index; WC, waist circumference; SBP, systolic blood pressure; DBP, diastolic blood pressure; HOMA-IR, homeostasis model assessment for insulin resistance; QUICKI, quantitative insulin sensitivity check index; TAG, triglycerides; HDL-c, high-density lipoproteins-cholesterol;; hsCRP, high-sensitivity CRP; MCP-1,monocyte chemoattractant protein 1; GGT, Gammaglutamyltransferase; TNF-α, tumour necrosis factor alpha; TSH, thyroid-stimulating hormone; IL, interleukin; PAI-1, plasminogen activator inhibitor-1; MPO, myeloperoxidase; sICAM, soluble intercellular cell adhesion molecule-1. *P<0.05.

| Outcome | Beta | SE | CI.LOW | CI.HIGH | T-value | P-Value | FDR |
| --- | --- | --- | --- | --- | --- | --- | --- |
| BMI Z-score | 0.356742 | 0.018365 | 0.017981 | 0.089973 | 2.939048 | 0.005009333* | 0.1302427 |
| MPO (µg/L) | -0.34989 | 2.845957 | -11.495421 | -0.339269 | -2.079211 | 0.043596686* | 0.4932982 |
| SBP | 0.262452 | 0.2514465 | -0.04712895 | 0.9385412 | 1.772,569 | 0.08292251 | 0.4932982 |
| AST (U/L) | 0.273129 | 0.066067 | -0.017778 | 0.241207 | 1.690916 | 0.097474922 | 0.4932982 |
| ALT (U/L) | 0.235236 | 0.113399 | -0.033886 | 0.410636 | 1.661178 | 0.103196554 | 0.4932982 |
| HsCRP (mg/L) | 0.234346 | 0.348906 | -0.128352 | 1.239357 | 1.592128 | 0.118205328 | 0.4932982 |
| TAG (mg/dL) | 0.232391 | 0.404079 | -0.173903 | 1.410089 | 1.529632 | 0.132811041 | 0.4932982 |
| Resistin (µg/L) | 0.223976 | 0.104071 | -0.057597 | 0.350361 | 1.406563 | 0.165998677 | 0.5394957 |
| sICAM1 (mg/L) | -0.208464 | 0.009806 | -0.031567 | 0.006874 | -1.259028 | 0.214108936 | 0.6185369 |
| GGT (U/L) | 0.190581 | 0.104644 | -0.081142 | 0.329061 | 1.184585 | 0.242137353 | 0.6295571 |
| DBP | 0.1500147 | 0.1634918 | -0.1569578 | 0.48393 | 0.9999652 | 0.3225587 | 0.6671418 |
| MCP1 (ng/L) | 0.162466 | 0.694272 | -0.676507 | 2.045038 | 0.985587 | 0.329277816 | 0.6671418 |
| TNFα (ng/L) | 0.14881 | 0.012211 | -0.012852 | 0.035016 | 0.907526 | 0.368662495 | 0.6671418 |
| Leptin (µg/L) | 0.079798 | 0.057139 | -0.06128 | 0.162704 | 0.887529 | 0.379220108 | 0.6671418 |
| IL6 (ng/L) | -0.18923 | 0.788794 | -2.241604 | 0.850468 | -0.881812 | 0.384889528 | 0.6671418 |
| QUICKI | 0.122802 | 0.000445 | -0.00051 | 0.001233 | 0.813133 | 0.420075148 | 0.6826221 |
| tPAI1 (µg/L) | -0.082656 | 0.165824 | -0.427502 | 0.222527 | -0.618051 | 0.539463347 | 0.7869461 |
| P Selectine (µg/L) | -0.09031 | 0.292972 | -0.752906 | 0.395543 | -0.609894 | 0.544808842 | 0.7869461 |
| HDL-c (mg/dL) | -0.054999 | 0.162272 | -0.382981 | 0.253125 | -0.400119 | 0.690881956 | 0.8600189 |
| Hip Circumference (cm) | -0.026917 | 0.09899 | -0.229915 | 0.158124 | -0.362617 | 0.718483164 | 0.8600189 |
| IL8 (ng/L) | -0.056542 | 0.087295 | -0.200509 | 0.141689 | -0.336899 | 0.737662007 | 0.8600189 |
| Insulin (mU/L) | -0.044402 | 0.117725 | -0.266814 | 0.194668 | -0.306415 | 0.760587143 | 0.8600189 |
| HOMA-IR | -0.045483 | 0.026985 | -0.061153 | 0.04463 | -0.306152 | 0.760785993 | 0.8600189 |
| Adiponectin (mg/L) | 0.021981 | 0.103484 | -0.186343 | 0.219314 | 0.159307 | 0.874095447 | 0.9469367 |
| Glucose (mg/dL) | -0.001604 | 0.112144 | -0.221027 | 0.218578 | -0.010919 | 0.99133255 | 0.9913325 |

**Table 2.** Association between the change in ISM1 serum levels and the change in metabolic outcomes in girls in the cross-sectional population (PUBMEP study). Multiple regression analyses with the change in ISM1 serum levels as independent variable. Models were adjusted for the change in BMI *Z*-Score, the origin, and the pubertal stage reached. When the dependent variable was the change in the BMI *Z*-Score, we replaced the BMI confounder with the change in Insulin levels. The change in Height was further included in models when the dependent variable was change in Blood pressure. Abbreviations: SE, standard error; CI, confidence interval; AST, aspartate aminotransferase; ALT, alanine aminotransferase; BMI, body mass index; WC, waist circumference; SBP, systolic blood pressure; DBP, diastolic blood pressure; HOMA-IR, homeostasis model assessment for insulin resistance; QUICKI, quantitative insulin sensitivity check index; TAG, triacylglycerols; HDL-c, high-density lipoproteins-cholesterol; hsCRP, high-sensitivity CRP; MCP-1,monocyte chemoattractant protein 1; GGT, Gammaglutamyltransferase; TNF-α, tumour necrosis factor alpha; TSH, thyroid-stimulating hormone; IL, interleukin; PAI-1, plasminogen activator inhibitor-1; MPO, myeloperoxidase; sICAM, soluble intercellular cell adhesion molecule-1. *P<0.05.

| Outcome | Beta | SE | CI.LOW | CI.HIGH | T-value | P-Value | FDR |
| --- | --- | --- | --- | --- | --- | --- | --- |
| HsCRP (mg/L) | -0.212532 | 0.275906 | -0.993822 | 0.087729 | -1.642031 | 0.10618814 | 0.9518221 |
| tPAI1 (µg/L) | -0.203555 | 0.222168 | -0.796221 | 0.074676 | -1.623874 | 0.109825622 | 0.9518221 |
| AST (U/L) | -0.161062 | 0.089849 | -0.2849 | 0.067309 | -1.210869 | 0.230856581 | 0.9591424 |
| BMI Z-score | 0.131089 | 0.024629 | -0.02142 | 0.075127 | 1.090312 | 0.280008946 | 0.9591424 |
| Adiponectin (mg/L) | -0.14196 | 0.115312 | -0.350002 | 0.102022 | -1.075251 | 0.286714995 | 0.9591424 |
| ALT (U/L) | -0.122581 | 0.084927 | -0.254045 | 0.07887 | -1.031329 | 0.306668253 | 0.9591424 |
| GGT (U/L) | -0.122033 | 0.068911 | -0.199177 | 0.070953 | -0.930365 | 0.35603822 | 0.9591424 |
| QUICKI | -0.091113 | 0.00047 | -0.001278 | 0.000563 | -0.760771 | 0.449823868 | 0.9591424 |
| HDL-c (mg/dL) | 0.094496 | 0.228087 | -0.278695 | 0.615407 | 0.738122 | 0.463416645 | 0.9591424 |
| HOMA-IR | 0.087244 | 0.034593 | -0.042877 | 0.092727 | 0.720527 | 0.474046011 | 0.9591424 |
| Insulin (mU/L) | 0.075117 | 0.144802 | -0.191328 | 0.376296 | 0.638694 | 0.525493807 | 0.9591424 |
| Resistin (µg/L) | 0.088929 | 0.20056 | -0.267462 | 0.518732 | 0.626421 | 0.533496578 | 0.9591424 |
| DBP | 0.1065075 | 0.1772502 | -0.2105193 | 0.4843014 | 0.772304 | 0.4433014 | 0.9591424 |
| MPO (µg/L) | 0.072895 | 2.284379 | -3.32945 | 5.625316 | 0.502514 | 0.617556761 | 0.9591424 |
| IL6 (ng/L) | 0.076885 | 0.679751 | -1.022602 | 1.642022 | 0.455622 | 0.651008589 | 0.9591424 |
| TNFα (ng/L) | -0.05857 | 0.014978 | -0.035842 | 0.022873 | -0.432909 | 0.666686377 | 0.9591424 |
| Glucose (mg/dL) | 0.048341 | 0.153445 | -0.247778 | 0.353727 | 0.345234 | 0.731167604 | 0.9591424 |
| sICAM1 (mg/L) | -0.037412 | 0.005192 | -0.011521 | 0.008831 | -0.258988 | 0.796561463 | 0.9591424 |
| Leptin (µg/L) | -0.027564 | 0.119642 | -0.264583 | 0.204416 | -0.251446 | 0.802358281 | 0.9591424 |
| SBP | 0.04020589 | 0.174974 | -0.3043049 | 0.4120564 | 0.2948135 | 0.7692668 | 0.9591424 |
| TAG (mg/dL) | 0.018997 | 0.597412 | -1.083729 | 1.258128 | 0.145962 | 0.884457754 | 0.9591424 |
| IL8 (ng/L) | -0.020783 | 0.07327 | -0.154252 | 0.132967 | -0.145253 | 0.885014516 | 0.9591424 |
| Hip Circumference (cm) | -0.004426 | 0.094848 | -0.193113 | 0.178692 | -0.076024 | 0.939666438 | 0.9591424 |
| MCP1 (ng/L) | -0.008154 | 0.724891 | -1.461766 | 1.379808 | -0.056531 | 0.955113059 | 0.9591424 |
| P Selectine (µg/L) | -0.006652 | 0.221513 | -0.445564 | 0.422768 | -0.051456 | 0.959142396 | 0.9591424 |

**Figure 1.**
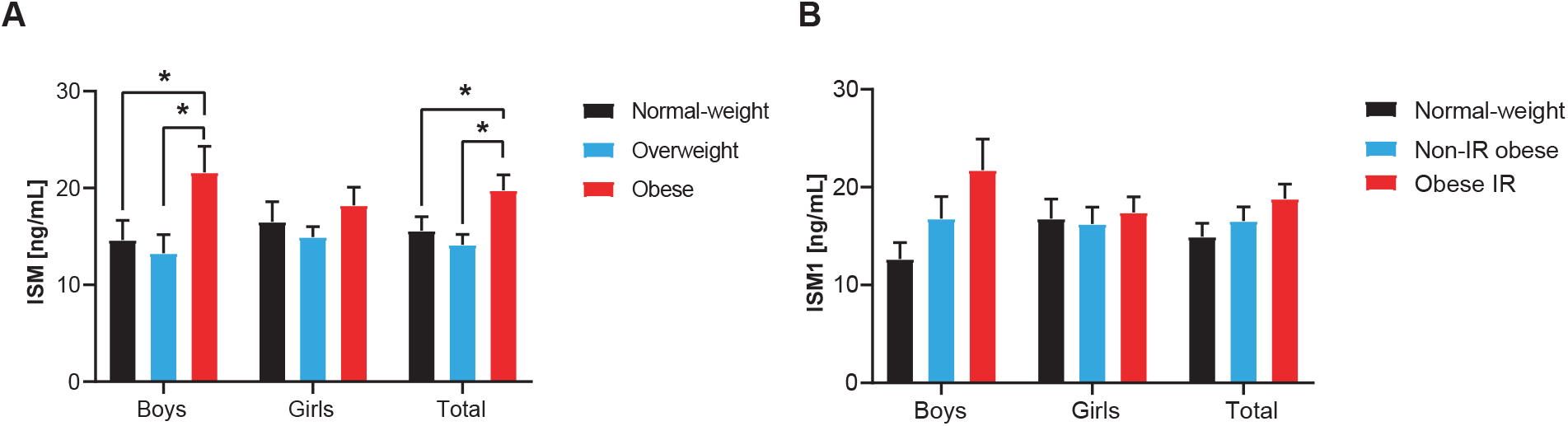
Group comparisons for ISM1 serum levels (ng/mL) in the pubertal population of 119 children. **A**) Comparison between normal-weight, overweight and obese. **B**) Comparison between normal-weight non-IR, non-IR and IR in children. The two-way ANOVA, Tukey’s multiple comparisons test was employed to assess group differences in ISM1 levels according to standard statistical assumptions. *P<0.05

**Figure 2.**
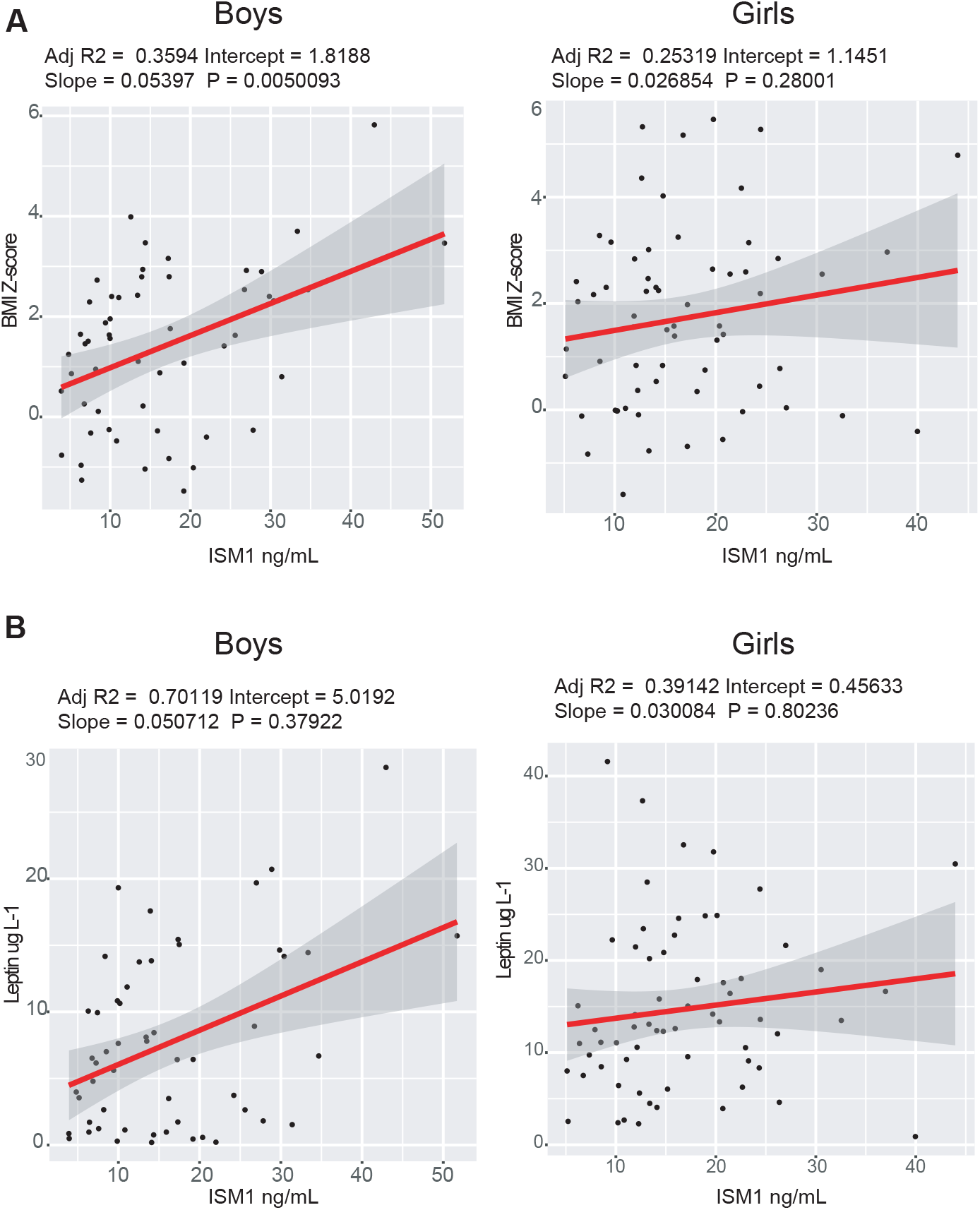
Multiple linear regressions analyses between the change in ISM1 serum levels (ng/mL) and the changes in BMI Z-score and leptin (µg/L) in the cross-sectional cohort. **A** reports the linear model with delta BMI Z-score as dependent variable, **B** refers to the model for delta leptin as dependent variable.

**Figure 3.**
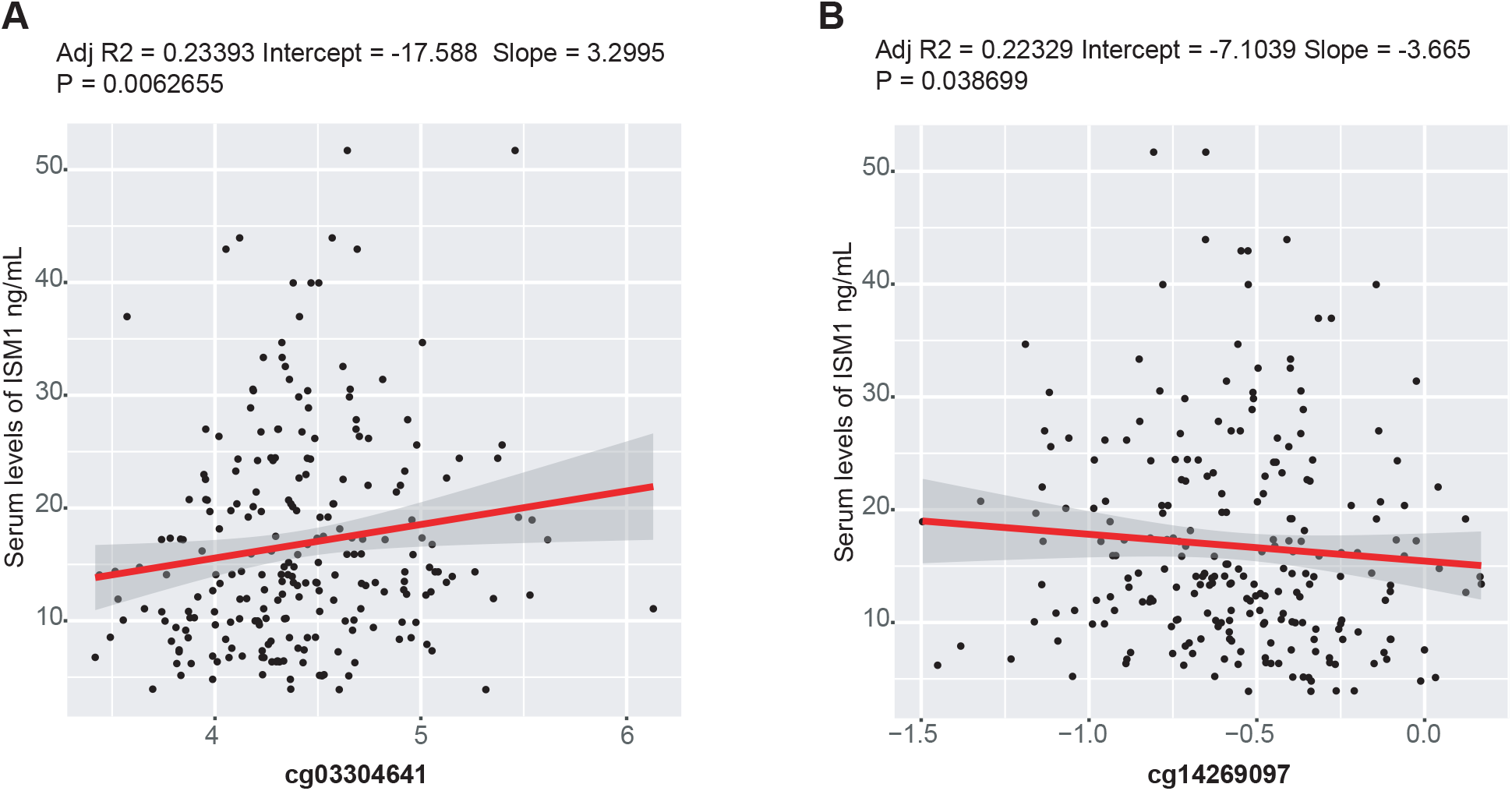
Multiple linear regressions analyses between the change in ISM1DNAmethylation status and ISM1 serum levels (ng/mL). **A** reports the linear model with delta ISM1 serum levels as dependent variable and cg03304641 as independent variable. **B** refers the linear model with delta ISM1 serum levels as dependent variable and cg14269097 as independent variable.

## Discussion

In the present work, we show higher serum levels of the novel insulin-like adipokine ISM1 in pubertal children with obesity, with a strong association with BMI *Z*-score in boys. Moreover, ISM1 was also associated with MPO, an enzyme associated with IR and inflammation in individuals with overweight [15]. These findings illustrate how this protein might be of special relevance as a new biomarker of obesity in children.

In obesity, hypertrophic adipocytes and adipose tissue-resident immune cells accelerate a low-grade and chronic, proinflammatory profile with altered secretion of adipokines and lipokines, thereby exacerbating cardiometabolic disease [16]. In this context, the production and secretion of adipokines and lipokines, which contribute to systemic energy metabolism by different mechanisms, are dependent on the energy status of adipose tissue. Hence, further preclinical and clinical studies exhibiting the activation or inhibiting the signaling of specific adipokines or lipokines (e.g. by using adipokine-neutralizing antibodies) may be an approach suitable to treat or prevent the development of metabolic disease. Nevertheless, efficacy and safety in humans need to be confirmed [17]. ISM1 is a secreted protein, originally discovered in fetal brain development and expressed in the brain, lung, vasculature, skin and immune cells [18]. ISM1 has been recently identified in mouse and human adipocytes, regulating glucose uptake while suppressing hepatic lipid synthesis, thus improving hyperglycemia and reducing lipid accumulation in mouse models. Besides, circulating plasma levels of human ISM1 has been detected at an average of 50 pg/mL and trend to positively correlate with BMI, but not with glucose in female individuals [7]. Now, we reveal a relationship between circulating levels of ISM1 and obesity is investigated in a pubertal population. The motivation for focusing on the identification of new biomarkers in puberty lies in the fact that sexual maturation has been presented as a significant metabolic risk period for children with obesity [4], and indeed, we previously described the role of S100A4 in IR through a multi-omics approach in children, providing an interesting knowledge into the plausible molecular mechanism underlying that association [10].

Here, in a pubertal sample of 119 children, we show how higher levels of ISM1 in children with obesity compared to normal-weight and overweight, and a robust positive association between ISM1 serum levels and BMI *Z*-score in boys, indicating a putative sexual dimorphism. However, MPO was negatively associated with circulating levels of ISM1. Indeed, MPO is the most abundant protein in human neutrophils, playing a major role in inflammation, oxidative stress, lipoprotein oxidation, and atherosclerosis [15, 19]. Moreover, MPO deficient mice are resistant to diet-induced obesity and IR, and inhibition of MPO activity in the neutrophils decreases diet-induced IR in obese mice, and activation of MPO is associated with the development of obesity and obesity-associated IR [20]. Group comparisons for ISM1 levels also revealed significant results in boys, although no significant changes were observed when comparing extreme experimental conditions in relation to IR (normal-weight vs. obese with IR). Jiang *et al*. (2021) [7] determined that ISM1 signaling is dependent on PI3K and shares downstream phosphorylation targets with insulin signaling, such as -AKT, p-AKT, p-ERK1/2 and p-S6. Outstandingly, ISM1 activates a PI3K-AKT signaling pathway independently of the insulin and insulin-like growth factor 1 receptors, being most likely to signal through another, yet to be identified, receptor tyrosine kinase. We found a correlation with obesity but not with HOMA-IR in children, which would point to the adipokine ISM1 with a direct role in obesity but not in the metabolic status derived from IR. Remarkably, while the glucoregulatory function of the novel adipokine is shared with insulin, ISM1 also neutralizes lipid accumulation in the liver by inhibiting *de novo* lipogenesis, promoting protein synthesis and preventing hepatic steatosis in a diet-induced fatty liver mouse model. Nevertheless, in addition to a disturbed hepatic and postprandial lipoprotein metabolism, enhanced triacylglycerol lipolysis in adipocytes and subsequent fatty acid flux to the liver are major determinants of hepatic steatosis [21]. As insulin is a major anti-lipolytic hormone in adipocytes, therefore, it is plausible that ISM1 signaling indirectly modulates hepatic lipid accumulation by inhibiting fatty acid release from adipose tissue in mice (Heeren 2021a). Additionally, in the paper of Jiang *et al*. (2021) [7], the therapeutic dosing of recombinant ISM1 improves glucose tolerance to the same degree as metformin, enhances diabetes in diet-induced obese mice and ameliorates hepatic steatosis in a diet-induced fatty liver mouse model, establishing that the recombinant Ism1 and its derivatives may be explored for therapeutic purposes and may offer certain advantages over current monotherapies. Nonetheless, the observed higher circulating ISM1 levels in pubertal children suggest that maybe an ISM1 resistance is present, as administration of ISM1 into mice with established disease improves glucose and lipid dysfunction in the diet-induced obesity. Additionally, we should consider that pubertal children did not show hyperglycemia, which it might mask the observed effects of ISM1 in mice.

The strengths of our findings are the relatively high number of recruited children from different centers in the country (Andalucia, Galicia and Aragón); the novelty of the recently described new adipokine, which has not been reported elsewhere in humans, and the possibility to correlate with several plasma adipokines, inflammation, and cardiovascular risk biomarkers in the subcohort. Furthermore, the availability of DNA methylation analysis by using the Infinium MethylationEPIC microarray using bead chip technology in all the children population allowed us to observe two-enhancer related CpG sites of ISM1 (cg03304641 and cg14269097) associated with serum levels of ISM1 in children. As a limitation, though ISM1 serum levels are associated with BMI *Z*-score, we cannot distinguish the origin of the circulating levels simply because ISM1 is secreted by other non-adipose cells, and a secondary validation population would be needed to confirm these findings.

In conclusion, the circulating levels of the novel insulin-like adipokine ISM1 are significantly higher in pubertal children with obesity, and strongly associated with BMI *Z*-score in boys. Furthermore, we reveal two DNA methylations in two-enhancer-related CpG sites of the ISM1 region associated with serum levels of the protein in children with obesity.

## Data Availability

All data produced in the present study are available upon reasonable request to the authors

## Acknowledgements

The authors would like to thank the children and parents who participated in the study.

## Author contributions

CAG is the guarantor of this work. AG and CAG contributed to the study concept of the cross-sectional study. FJRO, AAR and CAG design this study. RL, GB, MG, and LM participated in the child recruitment and anthropometric measures (data acquisition). CAG and AAR revised DNA extraction and methylation analyses. FJRO and MCR planned and performed the ISM1 analysis in serum of the children. AAR and FJRO performed all data analyses. AAR, FJRO and CAG took part in the interpretation of data. FJRO and CAG wrote the manuscript. All authors took park in the critical revision of the manuscript. CAG, RL and GB obtained funding. All authors approved the final version of the manuscript.

## Abbreviations

IR: insulin resistance
WC: waist circumference
TNF-α: tumour necrosis factor alpha
hsCRP: high-sensitivity CRP
IL: interleukin
PAI-1: total plasminogen activator inhibitor-1
MPO: myeloperoxidase
MCP-1: monocyte chemoattractant protein 1
MMP-9: matrix metalloproteinase-9
sICAM-1: soluble intercellular cell adhesion molecule-1
VCAM: soluble vascular cell adhesion molecule-1
MLR: multiple linear regression
FDR: false discovery rate
LME: linear mixed-effects
ISM1: isthmin-1
T2D: type 2 diabetes
MPO: myeloperoxidase.

## Supplementary Figures

**Figure S1 related to Figure 2.**
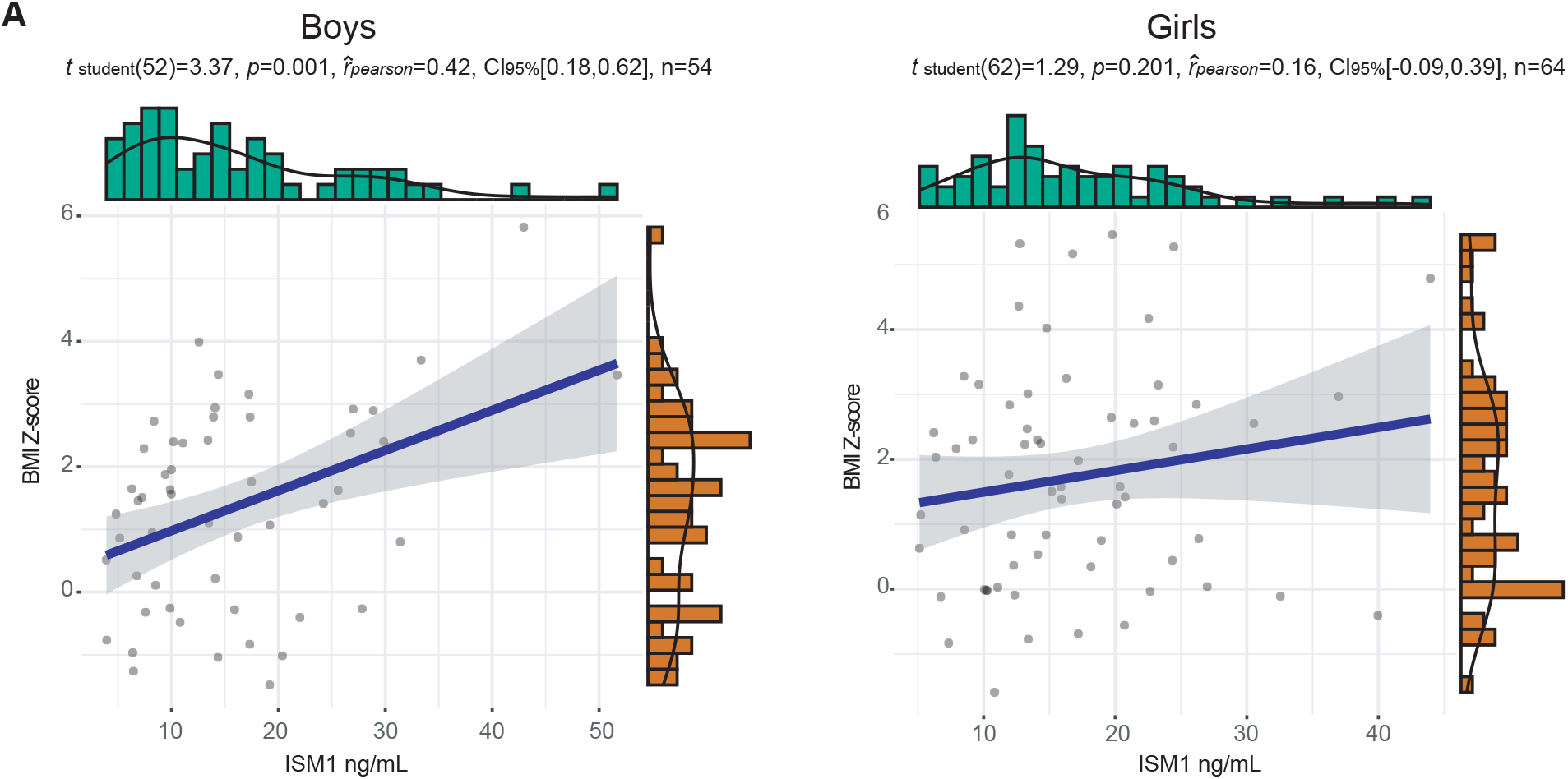
Multiple linear regressions were employed with M values as independent variables and each outcome as the dependent variable. Models were adjusted for confounders when necessary. Because percentage methylation is easily interpretable, beta values were employed for the graphical representation of results. **A** refers to BMI Z-score as dependent variable.

**Figure S2 related to Figure 3.**
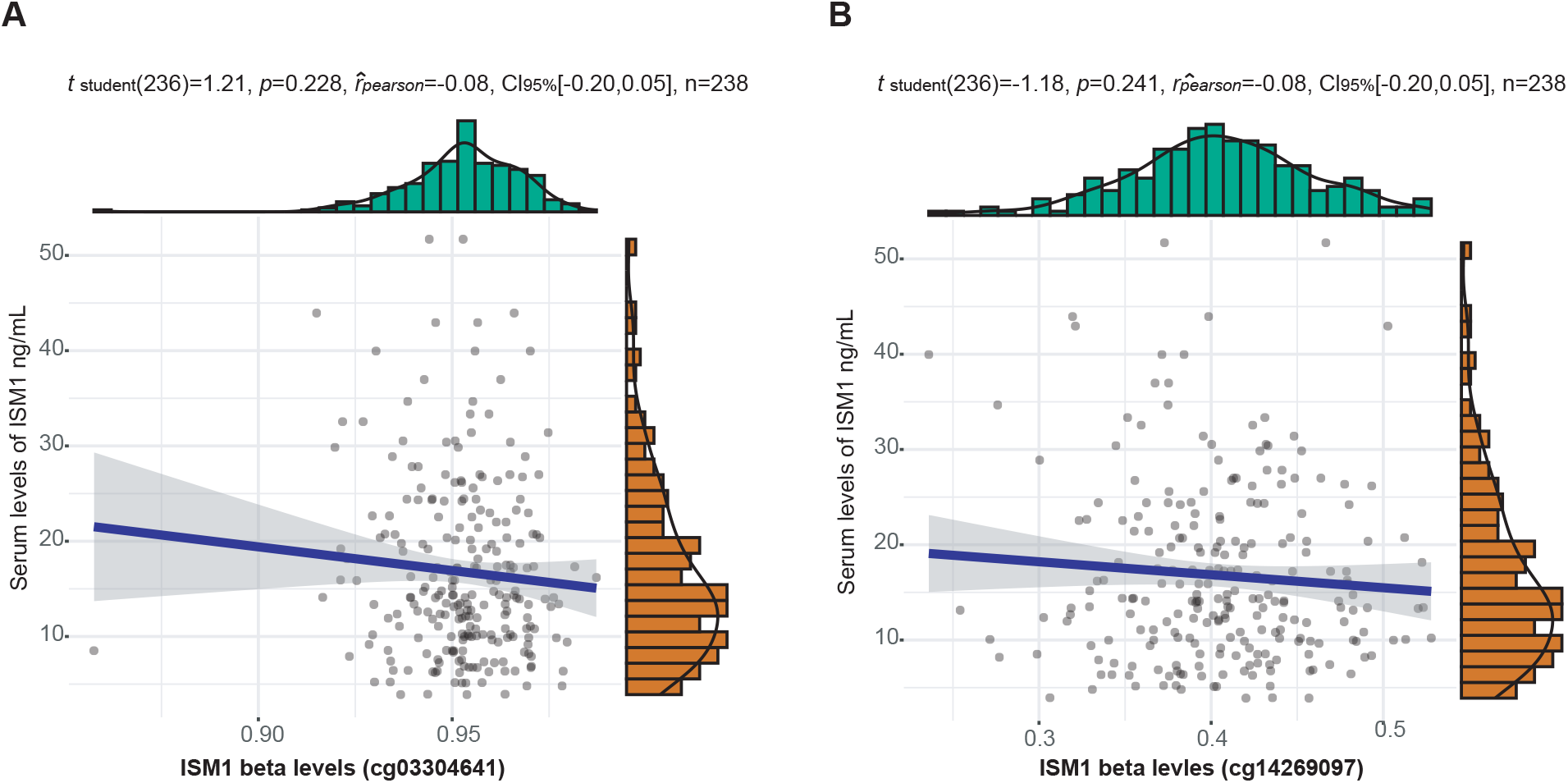
Cross-sectional associations between ISM1DNAmethylation status and ISM1 serum levels (ng/mL). Multiple linear regressions were employed with M values as independent variables and each outcome as the dependent variable. Models were adjusted for confounders when necessary. Because percentage methylation is easily interpretable, beta values were employed for the graphical representation of results.

**Supplementary table 1.** General characteristics, anthropometry, biochemical parameters, adipokines and cardiovascular/pro-inflammatory biomarkers in the cross-sectional cohort of 119 Spanish children.

|  | Normal-weight | Overweight | Obese | P-Value |
| --- | --- | --- | --- | --- |
| Age | 14.87 [11.22-17.59] NA=0 <sup>a</sup> | 14.22 [9.74-16.85] NA=0 <sup>a</sup> | 14.33 [9.72-18.07] NA=0 <sup>a</sup> | 0.15170492 |
| Sex | Girls=20; Boys=17 NA=0 | Girls=19; Boys=17 NA=0 | Girls=25; Boys=20 NA=0 | 0.97257376 |
| Glucose (mg/dL) | 85.11 (9.03) NA=0 <sup>a</sup> | 85.33 (6.76) NA=0 <sup>a</sup> | 85.2 (8.71) NA=0 <sup>a</sup> | 0.78038444 |
| TAG (mg/dL) | 63.25 (27.5) NA=1 <sup>b</sup> | 66.69 (31.29) NA=0 <sup>ab</sup> | 85.64 (37.49) NA=1 <sup>a</sup> | 0.00101011 |
| HDLc (mg/dL) | 56.5 [32-86] NA=1 <sup>a</sup> | 47 [26-70] NA=0 <sup>b</sup> | 43 [3-107] NA=1 <sup>b</sup> | 2.92E-06 |
| AST (U/L) | 22.36 (3.82) NA=1 <sup>a</sup> | 20.94 (6.27) NA=0 <sup>a</sup> | 22 (5.64) NA=1 <sup>a</sup> | 0.15850533 |
| ALT (U/L) | 13.62 (4.16) NA=0 <sup>b</sup> | 16.58 (6.34) NA=0 <sup>ab</sup> | 21.11 (8.61) NA=0 <sup>a</sup> | 1.22E-05 |
| GGT (U/L) | 11 [6-27] NA=1 <sup>a</sup> | 13 [1-27] NA=0 <sup>ab</sup> | 15 [2-44] NA=1 <sup>b</sup> | 0.00117436 |
| Insulin (mU/L) | 8.73 (3.52) NA=0 <sup>b</sup> | 12.58 (6.13) NA=0 <sup>b</sup> | 21.08 (11.52) NA=0 <sup>a</sup> | 6.60E-09 |
| HsCRP (mg/L) | 0.63 [0.2-30.1] NA=2 <sup>a</sup> | 1.78 [0.2-18.4] NA=2 <sup>b</sup> | 4.38 [0.2-164.2] NA=0 <sup>b</sup> | 2.89E-05 |
| Waist Hip ratio | 0.82 [0.54-1.38] NA=0 <sup>a</sup> | 0.83 [0.74-1] NA=1 <sup>a</sup> | 0.93 [0.75-2.11] NA=1 <sup>b</sup> | 8.42E-06 |
| WC Height ratio | 0.43 (0.05) NA=0 <sup>c</sup> | 0.52 (0.05) NA=1 <sup>b</sup> | 0.61 (0.08) NA=1 <sup>a</sup> | 1.47E-28 |
| QUICKI | 0.35 [0.32-0.44] NA=0 <sup>a</sup> | 0.34 [0.29-0.39] NA=0 <sup>b</sup> | 0.31 [0.28-0.42] NA=0 <sup>c</sup> | 7.70E-08 |
| DBP (mm Hg) | 67.16 (12.49) NA=2 <sup>a</sup> | 67.93 (8.11) NA=2 <sup>a</sup> | 70.44 (11.51) NA=1 <sup>a</sup> | 0.29095778 |
| SBP (mm Hg) | 105.77 (13.7) NA=2 <sup>b</sup> | 111.07 (12.35) NA=2 <sup>ab</sup> | 116.7 (15.42) NA=1 <sup>a</sup> | 0.01356016 |
| Glucose (mmol/L) | 4.72 (0.5) NA=0 <sup>a</sup> | 4.74 (0.38) NA=0 <sup>a</sup> | 4.73 (0.48) NA=0 <sup>a</sup> | 0.78038444 |
| HOMA-IR | 1.84 (0.78) NA=0 <sup>b</sup> | 2.69 (1.44) NA=0 <sup>b</sup> | 4.53 (2.71) NA=0 <sup>a</sup> | 5.38E-08 |
| Waist Circumference (cm<sup>2</sup>) | 70.15 (7.99) NA=0 <sup>c</sup> | 83.08 (7.8) NA=1 <sup>b</sup> | 99.43 (11.47) NA=1 <sup>a</sup> | 2.44E-25 |
| Hip Circumference (cm<sup>2</sup>) | 85.37 (9.44) NA=0 <sup>c</sup> | 97.59 (7.96) NA=1 <sup>b</sup> | 108.88 (10.51) NA=1 <sup>a</sup> | 7.87E-18 |
| BMI Z-score | -0.12 [-1.59-0.78] NA=0 <sup>a</sup> | 1.51 [0.75-2.3] NA=0 <sup>b</sup> | 2.9 [2.03-5.82] NA=0 <sup>c</sup> | 2.89E-22 |
| sICAM1 (mg/L) | 0.07 [0.04-3.31] NA=0 <sup>a</sup> | 0.1 [0.06-2.03] NA=0 <sup>b</sup> | 0.1 [0.06-1.72] NA=0 <sup>b</sup> | 0.00219134 |
| MPO (µg/ L) | 23.5 [0.8-196.36] NA=8 <sup>a</sup> | 33.63 [0.88-1077.71] NA=3 <sup>a</sup> | 32.47 [0.2-744.47] NA=2 <sup>a</sup> | 0.31811663 |
| P Selectin (µg/L) | 42.17 (12.27) NA=0 <sup>b</sup> | 47.1 (15.59) NA=0 <sup>ab</sup> | 56.13 (20.87) NA=1 <sup>a</sup> | 0.00630924 |
| Adiponectin (mg/L) | 13.47 (8.58) NA=0 <sup>a</sup> | 12.57 (7.62) NA=0 <sup>a</sup> | 8.35 (5.26) NA=0 <sup>b</sup> | 0.00722112 |
| Resistin (µg/L) | 18.12 [9.19-44.31] NA=0 <sup>a</sup> | 18.97 [7.82-56.01] NA=0 <sup>a</sup> | 21.18 [10.91-77.23] NA=0 <sup>a</sup> | 0.07726358 |
| tPAI1 (µg/L) | 8.95 (9.75) NA=0 <sup>b</sup> | 16.67 (11.35) NA=0 <sup>b</sup> | 25.25 (14.54) NA=0 <sup>a</sup> | 1.40E-06 |
| IL6 (ng/L) | 8.28 [0.16-173.01] NA=9<sup>a</sup> | 2.89 [0.03-223.84] NA=17<sup>a</sup> | 3.67 [0.12-178.99] NA=8<sup>a</sup> | 0.14984527 |
| IL8 (ng/L) | 2.58 [0.56-27.33] NA=0<sup>a</sup> | 2.01 [0.27-30.27] NA=0<sup>a</sup> | 2.29 [0.13-22.8] NA=0<sup>a</sup> | 0.37108233 |
| MCP1 (ng/L) | 110.82 (39.87) NA=0<sup>a</sup> | 100.26 (48.49) NA=0<sup>a</sup> | 107.32 (39.99) NA=0<sup>a</sup> | 0.55119851 |
| TNFα (ng/L) | 2.73 (0.76) NA=0<sup>a</sup> | 2.57 (1.02) NA=0<sup>a</sup> | 2.66 (0.78) NA=0<sup>a</sup> | 0.71217629 |
| Leptin (ng/L) | 3.93 [0.19-21.63] NA=0<sup>a</sup> | 10.7 [1.53-41.58] NA=0<sup>b</sup> | 14.64 [6.42-37.32] NA=0<sup>c</sup> | 2.87E-10 |
| ISM1 (ng/mL) | 12.36 [3.9-32.55] NA=0<sup>a</sup> | 13.79 [4.81-31.4] NA=0<sup>a</sup> | 16.77 [6.21-51.7] NA=0<sup>a</sup> | 0.06447697 |

Data are expressed as mean (standard deviation) or median [min-max] if not normally distributed. One-way ANOVA, Kruskal-Wallis and the Welch test were employed to assess group differences. Distributions within the same row with unlike superscript letters were significantly different ( $p < 0.05$ ) according to Pairwise-t-tests, pairwise Mann–Whitney U-tests and Dunn tests. Childhood obesity was defined according to Cole et al. (2000). Abbreviations: ALR, adiponectin leptin ratio; Apo B, apolipoprotein B; Apo A, apolipoprotein A; BMI, body mass index; DBP, diastolic blood pressure; HC, hip circumference; HDL-c, high-density lipoproteins-cholesterol; HOMA-IR, homeostasis model assessment for insulin resistance; hsCRP, high-sensitivity C reactive protein; IL, interleukin; IR, insulin resistance; ISM1, isthmin-1; LDL-c, low-density lipoproteins-cholesterol; MCP-1, monocyte chemoattractant protein 1; MPO, myeloperoxidase; OB, obesity; PAI-1, plasminogen activator inhibitor-1; QUICKI, quantitative insulin sensitivity check index; SBP, systolic blood pressure; sICAM, soluble intercellular cell adhesion molecule-1; TAG, triacylglycerols; THS; Thyroid-Stimulating Hormone; TNF-α, tumour necrosis factor alpha; WC, waist circumference. NA: non-available value.

**Supplementary table 2.**
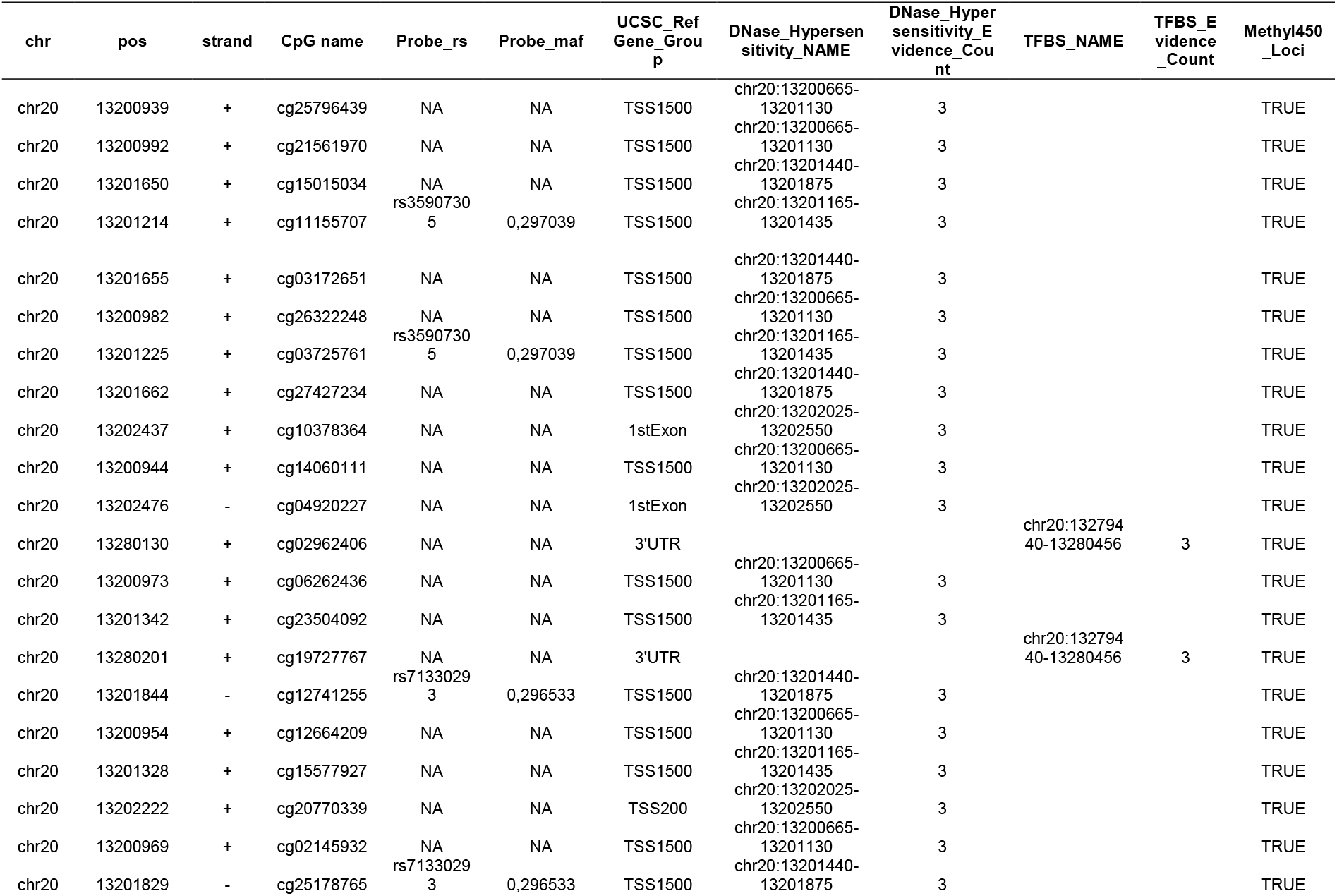
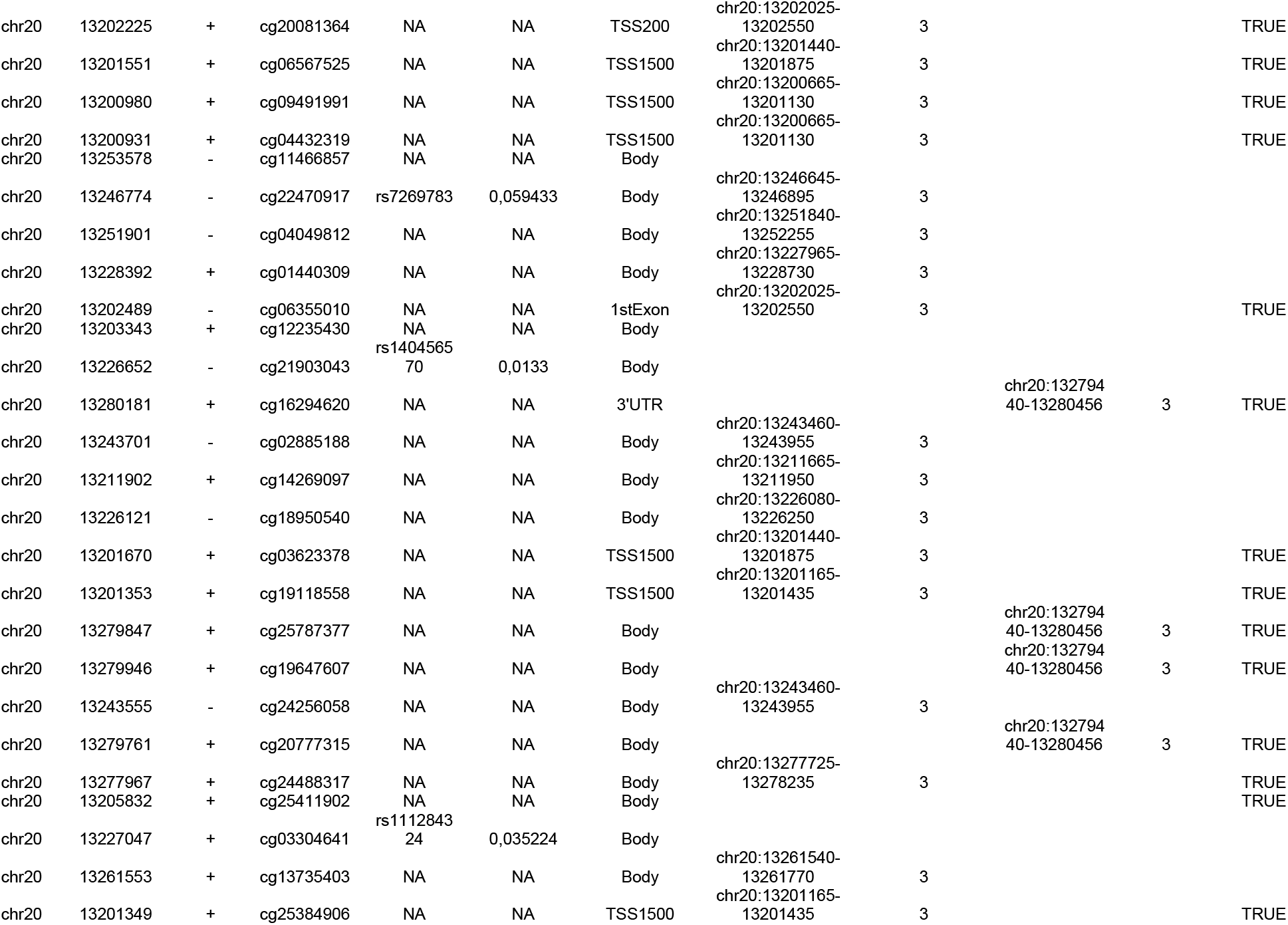
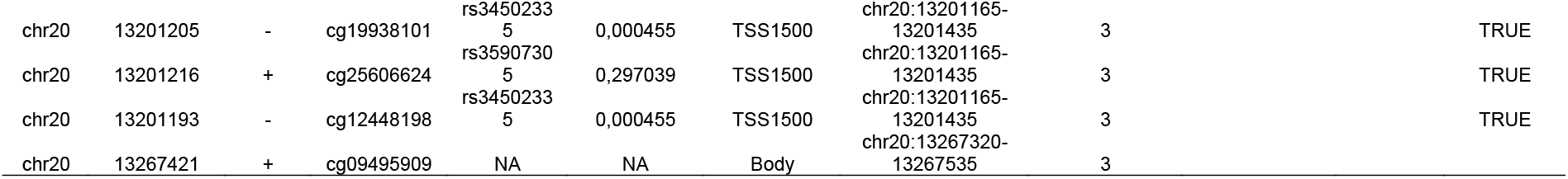
List of Mapped CpG sites in the ISM1 domain.

